# Moving forward through consensus: a national Delphi approach to determine the top research priorities in prostate cancer in Uganda

**DOI:** 10.1101/2023.05.17.23290089

**Authors:** Andrew Sentoogo Ssemata, Richard Muhumuza, Janet Seeley, Dorothy Chilambe Lombe, Monde Mwamba, Susan Msadabwe, Amos Deogratius Mwaka, Ajay Aggarwal

**Affiliations:** The Medical Research Council/ Uganda Virus Research Institute and London School of Hygiene & Tropical Medicine (MRC/UVRI & LSHTM) Uganda Research Unit, Entebbe, Uganda; Department of Global Health & Development, London School of Hygiene and Tropical Medicine, London, UK; Radiation Oncology, Te Whatu Ora Health NZ MidCentral, Palmerston North, New Zealand; ZAMBART, Lusaka, Zambia; Department of Radiation Oncology, Cancer Diseases Hospital, Lusaka, Zambia; Department of Medicine, College of Health Sciences, Makerere University, Kampala, Uganda; Department of Medicine, Faculty of Medicine, Gulu University, Gulu, Uganda; Department of Health Services Research and Policy, London School of Hygiene and Tropical Medicine, London, UK; King’s College London, London, UK

**Keywords:** Prostate cancer, delays, health care, Uganda, Delphi technique

## Abstract

**Objective:** To describe the health system, socioeconomic and clinical factors that influence access to and outcomes of care for prostate cancer (PC) in Uganda.

**Design:** Qualitative Delphi technique

**Setting:** Government and private-not-for-profit hospitals

**Methods:** We applied a two-stage modified Delphi technique to identify the consensus view across cancer experts. In the first round, experts received a questionnaire containing 22 statements drawn from a systematic review of the literature focusing on identifying the reason for the delay in accessing cancer care. Round 2 comprised 18 statements considered the greatest priority from round 1.

**Results:** We found that the top five research priority areas arise from challenges including: (i) lack of Diagnostic services - Ultrasound, laboratory tests, and biopsy facilities; (ii) High costs of services e.g., surgery, radiotherapy, hormone therapy are unaffordable to most patients, (iii) Lack of critical surgical supplies and essential/critical medicines, (iv) Lack of awareness of cancer as a disease and low recognition of symptoms, (v) Low Healthcare literacy. The lack of critical surgical supplies, high costs of diagnostic investigations and treatments were ranked highest in order of importance in round one. Round two of the survey also revealed lack of diagnostic services, unavailability of critical medicines, lack of radiotherapy options, high costs of treatments and lack of critical surgical supplies as the top priorities.

**Conclusion:** These research priority areas ought to be addressed in future research to improve prompt prostate cancer diagnosis and care in Uganda. There is need to improve the supply of high-quality affordable anticancer medicines provided timely to prostate cancer patients so as to improve the survivorship from the cancer.

**Article Summary:** *Strengths and limitations of this study:*

- This is a novel Delphi study exploring research priorities for prostate cancer research in Uganda from various stakeholders in cancer care.
- The study was informed by a recent systematic review which provided insight into the statements explored in the Delphi technique.
- Participants were recruited from various clinical, government aided and private settings and geographical locations giving representativeness of the participants.
- A limitation to this study was the low response rates in round 1 and round 2 as most stakeholders were not familiar with the Delphi technique, or how it works.

## Introduction

In 2020, there were 19.3 million new cancer cases and 10 million cancer deaths worldwide.^1^ Prostate cancer is among the top 3 most common incident cancers in men (1.3 million incident cases) and the leading cause of cancer-related deaths for men in 56 countries including Uganda.^2^

A recent review has presented a worrying disparity in the mortality to incidence ratio from prostate cancer at 90% in Africa compared to 10% in North America.^3^ For a curable cancer these figures are stark. In the low- and middle-income countries (LMICs), one of the challenges facing cancer control is the late diagnosis with advanced stage cancers at diagnosis.^4-6^ The reasons that contribute to delayed diagnoses and advanced stage at presentation include delayed recognition of symptoms and signs of cancers by patients and primary healthcare professionals, inadequate health and cancer information for patients, poor continuity of care, lack of facilities for biopsy and histological diagnoses, costs and availability for treatments including drug therapies.^6-8^

One of the key challenges to delivering timely prostate cancer care is that the condition is largely asymptomatic in its early stages. African men also face much higher prostate cancer frequency, severity, earlier age of disease and greater risk of death than is seen in Caucasian populations, leading to much greater economic impacts on societies.^9^

In Uganda, there has been much emphasis on cancers affecting women, especially breast and cervical cancers.^10-12^ Little attention has been given to the cancers affecting men especially prostate cancer that has been increasing in incidence over the recent decades.^13-15^ This apparent neglect and lack of data for planning for cancers affecting men has generated the need to prioritise key areas for research.

We present findings from the African Prostate Cancer - Disparities in Outcomes in Uganda (APC-DOUG) study whose aim was to describe health system, socioeconomic and clinical factors that influence access to and outcomes of care for prostate cancer across hospitals in Uganda. The purpose of this Delphi process was to prioritise key areas for research in prostate cancer in the Ugandan context by establishing the major barriers to seeking cancer care, reaching cancer care, and receiving high quality cancer care. By defining future research priorities and investigations, this will help to support the future development of health system interventions to reduce inequalities and improve prostate cancer outcomes in hospitals across Uganda.

## Methods

The Delphi technique is a relevant source of evidence in health care research foundational in the formation of consensus or the exploration of a field beyond existing knowledge.^16^ It has been used in medicine to develop research priorities.^16-18^

A structured 2-round modified Delphi technique with Ugandan clinical and health system research experts involved in prostate cancer care was conducted between December 2021 and July 2022. The Delphi technique was used to elicit consensus on the key research priorities in relation to the delays that may occur in receiving a diagnosis and/or treatment based on the Three Delays framework.^19 20^ The “Three Delays” Framework has been effective as a rapid health system assessment tool to understand complex multi-factorial problems affecting access to care.^20^ The Three delays framework focused on delays to: (i) Seeking care - delays in recognizing illness and deciding to seek appropriate medical help outside the home; (ii) Reaching care - delays in reaching an appropriate health facility and (iii) Receiving quality care - delays in receiving quality care after reaching the health facility.^21^

### Development of the survey

The Delphi survey was informed by two systematic reviews undertaken by the study team appraising primary quantitative,^15^ and qualitative (Mwamba et al., under peer review) research studies that had sought to identify the factors influencing delays in cancer diagnosis and treatment as well as receiving quality care in Sub-Saharan Africa. From this literature review 22 key factors contributing to delays were identified and represented priority areas for further investigation. These were translated into written statements that formed part of the Delphi consensus exercise for prioritizing research in prostate cancer in Uganda (supplemental appendix 1).

### Data collection

Data collection for round 1 of the Delphi process was conducted between December 2021 and February 2022 and round 2 of the Delphi process between March and July 2022.

Participants were selected to ensure representation from different regions, health care sectors (primary and secondary); specialist disciplines (urology, oncology); academia and public health as well as patient representatives. Lists of participants to approach from both public and private health facilities in Kampala, Wakiso, and Mukono districts were developed by the planning committee which comprised two oncologists, and two health services research methodological experts who oversaw the design, execution, and analysis of all phases of the study.

At present there is only one major comprehensive cancer treatment centre in Uganda (the Uganda Cancer Institute, based at Mulago Hospital) with two active regional bases (in Mbarara, western Uganda, and Gulu; Northern Uganda),^6^ and representation from each of these were sought. A snowballing approach was also undertaken where participants recommended additional stakeholders for participation. Agreement was reached regarding participant selection, consensus thresholds, and survey format and question structure. For round 1, we approached participants from the Uganda Cancer Institute, government hospitals, Private-for-profit hospitals, Private-not-for-profit hospitals, Ministry of Health, and a university.

### Procedure

Experienced social science and clinical researchers conducted the data collection using a survey questionnaire. Participants were approached by a formal invitation to participate by email correspondence followed by a phone call and for some participants a face-to-face meeting. For those willing to participate, we collected demographic data, and a structured questionnaire was hand delivered to the participants. This was followed up with a reminder phone call within a four-week period to ensure completion of the question. Two sequential rounds of questionnaires were used for this Delphi process (Supplemental appendix 1 and 2).

Round 1 consisted of scoring the 22 statements using a 4-point scale. Each statement was assigned a score (out of 20) based on the four criteria, each scored out of 5, as below:

- **Feasible** – how easy is it to measure or assess this reason for the delay?
- **Large scale** – does this factor affect a considerable proportion of men with prostate cancer.
- **High Impact** –is this factor a significant cause of death or disability from prostate cancer?
- **Modifiable** – can this factor be reasonably addressed to improve the care of men with prostate cancer.

Once all the questionnaires for round 1 were received, we compiled the rankings for each statement. Based on the criteria by Schneider et al,^22^ statements were selected as follows. Statements where 70% of participants gave a score of ≥15 were included as research priorities. Statements for which <30% of participants gave a score of ≥15 were excluded at this stage.^22^ The rest of the statements went forward to the second round of the Delphi consensus process. The information provided in the round 1 of the Delphi technique was collated and summarized in REDCap software,^23^ to enable formal analysis, and to formulate a second questionnaire with fewer statements based on the selection criteria above.

Round 2 of the Delphi included 18 statements. An electronic questionnaire designed in REDCap was generated and emailed to 42 participants. Participants were followed up with a reminder phone call within a four-week period and through face-to-face meetings by the study team. Recurrent emails and phone calls were made to those who had been contacted and had not completed the survey.

### Ethical considerations

This study was performed in line with the principles of the Declaration of Helsinki. Ethical approval to conduct the Delphi technique was obtained from the Uganda Virus Research Institute Research Ethics Committee (UVRI-REC) Ref: GC/127/21/09/859 and the Uganda National Council for Science and Technology (UNCST) Ref: HS1790ES and the London School of Hygiene and Tropical Medicine Ref: 26672. Each participant provided written informed consent before any study procedures were conducted and were reimbursed for their time and participation in the study. Each participant was provided a time compensation of 30,000 Uganda shillings at the end of each completed questionnaire.

## Results

Ten and twelve respondents completed the survey questionnaires in round 1 and 2 respectively. In round 1, a total of 30 clinicians were invited through email and face to face meetings, of which 10 (34%) participated and completed the survey. The majority of the participants in this round were oncologists *n* = 4 (40%) and cancer researchers *n* = 3 (30%). Most of the participants worked in a cancer care facility. In round 2, a total of 42 clinicians were invited through email, online and face to face meetings, of which 12 (29%) participated and completed the survey. Most of the participants in this round were radiologists *n* = 3 (25%) and general practitioners *n* = 3 (25%). Most of the participants in round 2 operated from a general health care setting. Of the 22 people who participated in round 1 and 2, three participated in both rounds and seven participants participated in round 1 only. The characteristics of the respondents in Rounds 1 and 2 are presented in table 1.

**Table 1.** showing characteristics of participants in round 1 and 2 of the Delphi process.

|  | **ROUND 1** |  |  |
| --- | --- | --- | --- |
|  | **Gender** | **Profession** | **Organisation** |
| 1 | Male | Oncologist | Cancer Institute, Kampala |
| 2 | Male | Oncologist | Cancer Institute, Kampala |
| 3 | Male | Oncologist | Cancer Institute, Kampala |
| 4 | Male | Cancer researcher and administrator | Cancer Institute, Kampala |
| 5 | Male | Oncologist | Cancer Institute, Kampala |
| 6 | Male | Public health specialist and policy expert | University/ Ministry of Health, Kampala |
| 7 | Male | Medical doctor/ General practitioner | Government hospital, Kampala |
| 8 | Female | Researcher and Clinical officer | Government hospital, Kampala |
| 9 | Male | Urologist | Cancer Institute, Kampala |
| 10 | Female | Surgeon and Cancer researcher | Government hospital, Kampala |
| **ROUND 2** |  |  |  |
| 1 | Female | Urologist | Private for-profit hospital, Kampala |
| 2 | Female | Surgeon and Cancer researcher | Government hospital, Kampala |
| 3 | Female | Researcher and clinical officer | Government hospital, Kampala |
| 4 | Male | Medical doctor/ General practitioner | Government hospital, Kampala |
| 5 | Male | Urologist | Government hospital, Kampala |
| 6 | Male | Medical doctor/ General practitioner | Private not-for-profit hospital, Kampala |
| 7 | Female | Radiologist | Private not-for-profit hospital, Kampala |
| 8 | Female | Radiologist | Private not-for-profit hospital, Kampala |
| 9 | Male | Surgeon | Government hospital, Mukono |
| 10 | Male | Medical doctor/ General practitioner | Government hospital, Mukono |
| 11 | Male | Surgeon | Private for-profit hospital, Kampala |
| 12 | Male | Radiologist | Private not-for-profit hospital, Kampala |

Following the first round of the Delphi technique, we excluded four statements of research priorities from the second-round questionnaire where <30% of participants had given them a score less than 15. These statements were related to barriers such as staff motivation, inadequate training etc. Two of the barriers scored greater than 70% and were to be included within our final selection of research priorities. These were “*lack of awareness of cancer as a disease*” and “*poor health care literacy*” (table 2).

**Table 2:** Ranking of Round 1 questionnaire statements.

|  | **Statement** | **Ranking (%**<br>**of**<br>**participants**<br>**scoring**<br>**reason >=15)** |
| --- | --- | --- |
| 1 | Lack of awareness of cancer as a disease and recognition of symptoms | 90 |
| 2 | Poor Healthcare literacy (when, how and where to seek services) | 90 |
| 3 | Lack of critical surgical supplies | 70 |
| 4 | Cost of diagnostic investigations | 70 |
| 5 | Cost of treatments e.g., surgery, radiotherapy, hormone therapy | 70 |
| 6 | Lack of Workforce (basic numbers low and inadequate training of staff) | 70 |
| 7 | Accessibility of care (long distance/travel times to access specialist services) | 60 |
| 8 | Difficulties with healthcare coordination between regions and hospitals as patients referred for specialist investigation and treatment | 60 |
| 9 | Lack of availability of Essential medicines | 60 |
| 10 | Cost of accessing healthcare (e.g., cost of accommodation and transport needed to receive treatment from centralized services) | 60 |
| 11 | Misdiagnosis of cancer at lower system levels (e.g., primary care, district hospital) | 50 |
| 12 | Lack of Diagnostic services (X-ray, Ultrasound, labs (e.g., PSA testing, biopsy facilities) | 50 |
| 13 | Lack of Radiotherapy options (brachytherapy/ teletherapy) | 50 |
| 14 | Preference for traditional, complementary, and alternative medicine | 40 |
| 15 | Lack of trust in healthcare and patients' citizens' rights (perceived quality; attitudes of healthcare workers; previous bad experience e.g., patients being turned away or refusal to refer; adequate consent) | 40 |
| 16 | Personal and professional obligations (financial and social implications to the patient and their families of seeking care and undergoing treatment) | 40 |
| 17 | Lack of social capital to support cancer journey especially where patients must travel for care (relationships, support from family, friends, colleagues) | 40 |
| 18 | Patient Fitness and treatment toxicity | 40 |
| 19 | Stigma associated with cancer diagnosis or severe illness/ Fears and beliefs around cancer | 30 |
| 20 | Communication/ language barrier between health care staff and patients | 20 |
| 21 | Staff motivation and burnout | 20 |

In Round 2, four statements reached the consensus target (>70% of respondents scoring the statements >=15) as shown in table 3. These were: (i) Lack of diagnostic services (X-ray, Ultrasound, labs (e.g., PSA testing, biopsy facilities, (ii) Lack of availability of critical medicines, (iii) Lack of Radiotherapy options (brachytherapy/ teletherapy), and (iv) Cost of treatment e.g., surgery, radiotherapy, hormone therapy. In total, six research priorities were identified during the two round Delphi technique.

**Table 3:**
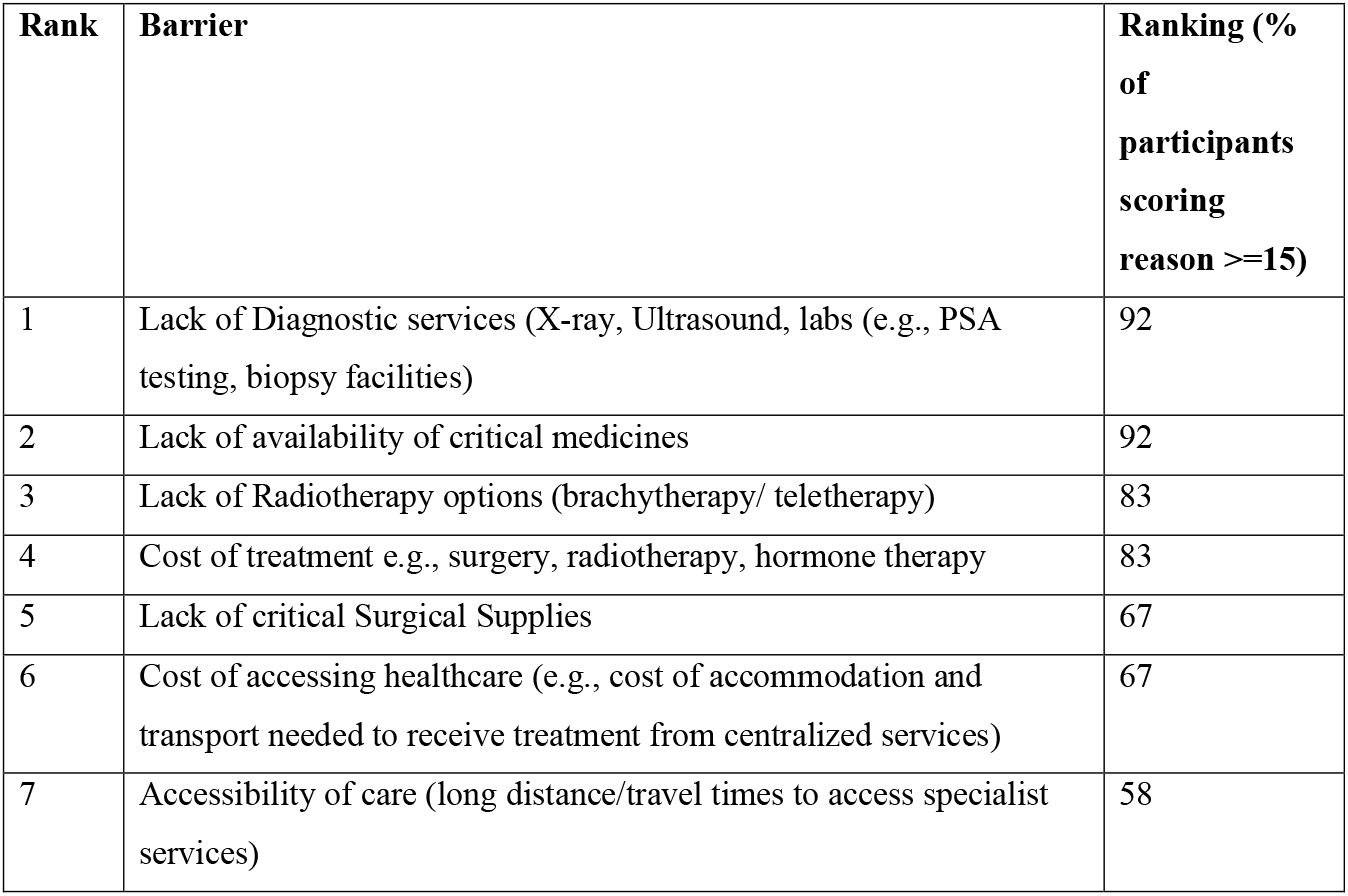
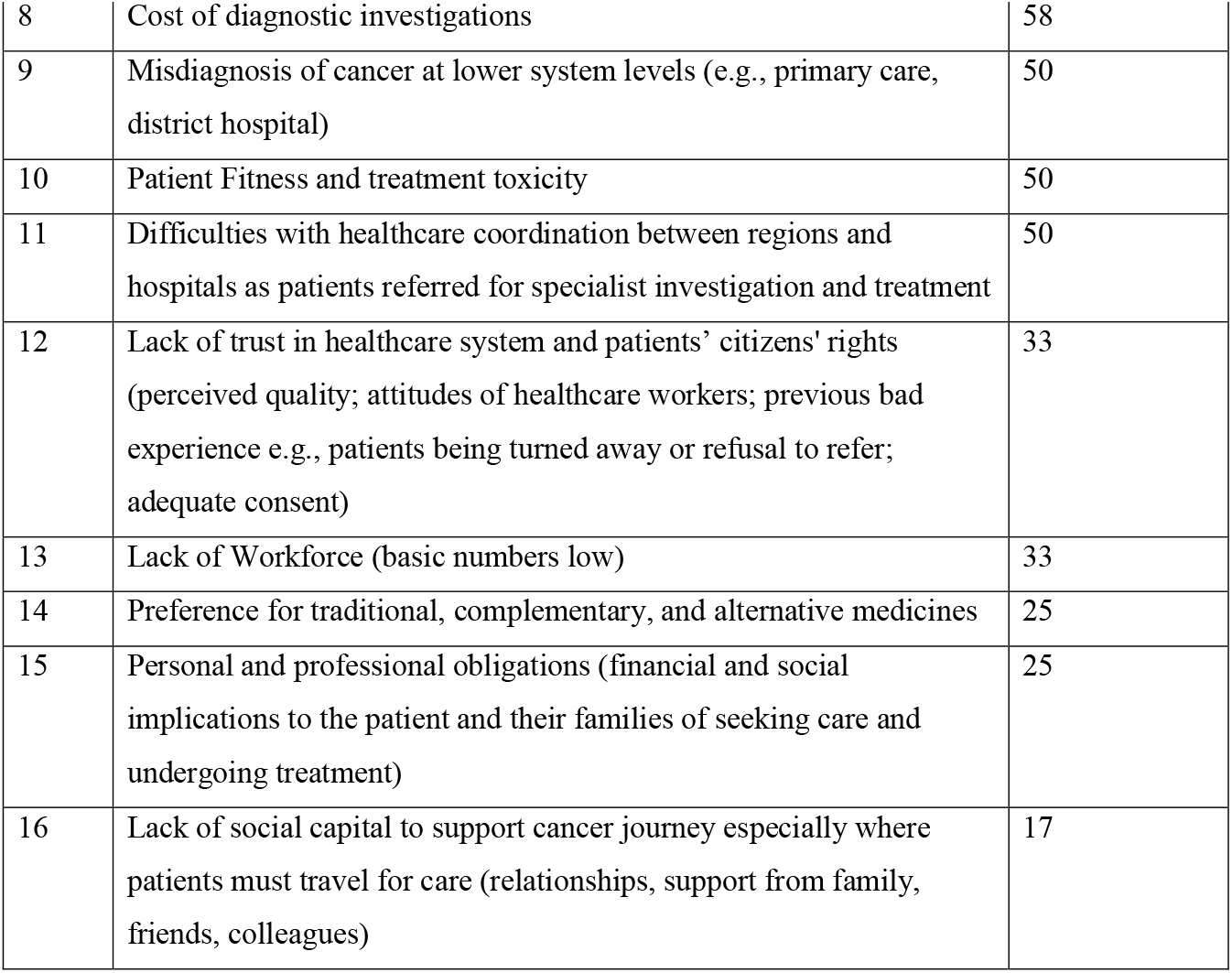
Round 2 ranking.

## Discussion

The purpose of this Delphi technique was to prioritise key areas for research in prostate cancer in the Ugandan context and will form the basis of recommendation for cancer control planning and research investments. The Delphi process sought to priorities those factors that had the greatest impact on achieving timely access to high quality care, could be investigated empirically and for which interventions could be developed and implemented.

Our study invited cancer experts, clinicians, and policy experts in and outside cancer management and care to evaluate the top research priorities for prostate cancer care in Uganda. Six key domains were identified: (i) lack of awareness of cancer as a disease and recognition of symptoms, (ii) poor healthcare literacy, (iii) lack of diagnostic services, (iv) lack of availability of essential medicines, (v) lack of radiotherapy facilities (vi) cost of treatment.

With respect to these six areas there remains a paucity of literature to date in the Ugandan context but are important issues as outlined below.

### Lack of awareness of cancer as a disease and poor recognition of cancer symptoms

Poor knowledge and misconceptions regarding prostate cancer and screening among Ugandan men,^24^ is likely to contribute to diagnosis with advanced clinical stages (stage III and IV) of PC.^25^ A lack of awareness about cancer and low health literacy has been reported as one of the major reasons for delays in seeking quality cancer care in sub-Saharan Africa.^15^ Similarly, the lack of awareness of prostate cancer its symptoms and its consequences, definitely affects the uptake of screening and seeking specialist help that provides an opportunity for early detection of prostate cancer. The lack of adequate information and counselling on prostate cancer has been associated with the considerably low uptake of prostate cancer screening among Ugandan men.^26^ Developing appropriate interventions for improving awareness and knowledge about prostate cancer are critical in supporting early detection and treatment outcomes of men who develop prostate cancer.

### Low health literacy (when, how and where to seek services)

Healthcare literacy is defined as the degree to which individuals have the capacity to obtain, process, and understand basic health information and services needed to make appropriate health decisions and to successfully navigate the health care system.^27, 28^ Poor healthcare literacy limits access to care, interaction with health services providers and illness management as people with low healthcare literacy are more likely to have poorer use of health services and therefore experience poorer health outcomes.^29^ A recent study among prostate cancer survivors in Uganda has shown that inconsistent information or complete lack of information are common experiences of the prostate cancer survivors can affect their health outcomes.^30 31^ Patients with limited health literacy may have limited knowledge and understanding of health and when coupled with lack of established follow-up mechanisms and inadequate social care support services experienced by many cancer patients in Uganda, this reduces their autonomy in self-care and decision making.^6^

### Lack of diagnostic services

A lack of basic imaging and pathological services results in delays in confirming a cancer diagnosis and completing staging to support management options. Such delays have a knock-on effect on the timely delivery of treatment leads to high mortality, morbidity, and low quality of life. ^25 32 33^ Lack of diagnostic services has led to delayed cancer diagnosis being a common in the Uganda setting.^31^ Additionally, where the service has been obtained the high costs of diagnostic investigations compounded by the poor social-economic status of the patients has resulted in enormous delays in seeking prostate cancer care among Ugandan men.^6^

### Lack of availability of essential medicines

A recent review has reported major barriers in access to core cancer medicines worldwide, with high prices are major barrier including medicine included in the WHO essential medicines list.^34^ Even relatively low cost medicines (compared to chemotherapy and molecularly targeted anti-cancer agents) such as Goserellin are largely unavailable to patients in the Ugandan setting due to recurrent stock-outs and majority being costly.^6^ A recent recommendation has suggested increased repurposing of existing drugs such as Metformin, Valproic acid initially intended for other conditions to treat prostate cancer.^35^

### Lack of Radiotherapy facilities

This is a core treatment for prostate cancer particularly in high-risk disease. However, radiotherapy is not commonly available in sub Saharan Africa with limited experience for radiotherapy and brachytherapy, insufficient infrastructure as well as limited trained personnel and training opportunities.^36^ Its low provision in Uganda has made the provision of treatment unachievable for many cancer sufferers. It has been previously noted that Uganda needs more than 20 operational radiotherapy units in order to respond adequately to its population demands.^37^ Currently the only radiotherapy machines in the country are all located at the UCI in the capital city Kampala. This has resulted in demand for services not being met (simply patients do not receive it), prolonged waiting times, compromises timing between the administration of radiation doses and eventually clinical and treatment outcomes. Even when potentially lifesaving radiotherapy treatment options are made available, there are challenges with the cost of treatment for a known cornerstone of curative therapy.

### The cost of treatment (surgery, radiotherapy, hormone therapy)

The cost of PC care is critical as there is limited access and availability to safe and reliable services including chemotherapy for prostate cancer patients in Uganda. The government of Uganda provides generic anticancer medicines for patients at the UCI at subsidised costs under the Universal Health Coverage (UHC) strategy to make available the WHO Essential anticancer Medicine List (EML).^38^ For example, at the UCI, patients pay a subsidised fee of approximately $85 before accessing radiotherapy services.^6^ In general, most cancer patients in Uganda are not able to afford cancer therapy out of pocket and yet the UHC arrangement does not make available newer anticancer therapies. The proportion of anticancer agents especially the newer targeted therapies on the 2019 WHO EML that are available on the 2016 Uganda National Essential Medicines List (NEML) was about 70.5%. Essential medicines for prostate cancer including Leuprolide, Bicalutamide and Abiraterone are often out of stock through the government UHC strategy.^39^ Availability and prices of anticancer agents vary widely in the LMICs including Uganda. The low availability and unaffordability of anticancer agents often lead patients to turn to alternative care approaches.^40^ PC is among the commonest cancers treated with complimentary therapies in Uganda. The unavailability of critical medicines and curative treatments being out of reach has pushed many patients to seek alternative, traditional, Chinese, and complementary medicines chiefly from potential anticancer medicinal plants.^6 41^

### Strengths and limitations

The Delphi technique was conducted by a multidisciplinary team following the CREDES guidelines.^16^ The statements explored in the Delphi were identified through a systematic review. We were able to use both electronic web-based as well as face-to-face paper-based approaches to data collection. This study has some limitations: the response rates in round 1 and round 2 were relatively low (34% and 29%, respectively). However, the aim of this study was to have suitable representation to identify the top research priorities for prostate cancer care in Uganda rather than receiving a high response rate. As the survey was only completed by clinical and health system research experts, majority of whom were not familiar with the Delphi technique, how it works, and this may have resulted in bias. Further research is required to elicit patients’ views. Although our questionnaires were sent to various groups and healthcare professionals from different clinical roles, most of the participants and experts in this study were oncologists and medical doctors. Future studies may need to consider other members of the healthcare team who were under-represented and may have alternative perspectives.

## Conclusion

This study has identified the top six research priorities for prostate cancer in Uganda. Our findings have implications for designing appropriate and contextual prostate care services locally to ensure men have fewer barriers to receiving earlier diagnosis and high-quality affordable treatment and survivorship care.

## Data Availability

All data produced in the present study are available upon reasonable request to the authors

## Acknowledgements

We thank the participants who participated in the Delphi technique and research team in Uganda where the study was conducted.

## Author contributions

AA conceived the study idea and designed it with AS, ADM and JS. AA, AS, AM, JS were engaged in the preparation and conduct of the Delphi study. AS, RM, DL, MM, SC, and AA led the writing of the paper and participated in the analysis. AA, ADM, and JS supervised the overall study. All authors read and approved the final manuscript.

## Declaration of conflicting interests

The author(s) declared no potential conflicts of interest with respect to the research, authorship, and/or publication of this article.

## Patient and public involvement

Patients and/or the public were not involved in the design, or conduct, or reporting, or dissemination plans of this research.

## Patient consent for publication

Not required.

## Ethics approval

Ethical approval was obtained from the Uganda Virus Research Institute Research Ethics Committee (UVRI-REC) Ref: GC/127/21/09/859 and the Uganda National Council for Science and Technology (UNCST) Ref: HS1790ES and the London School of Hygiene and Tropical Medicine Ref: 26672.

## Funding

The study was supported by funding from Wellcome’s Institutional Strategic Support Fund grant no. 204928/Z/16/Z through the London School of Hygiene and Tropical Medicine.

## Data sharing statement

Please contact the corresponding author in regard to any data sharing requests. Please note there are ethical restrictions on sharing of data.

## Supplemental Appendix 1

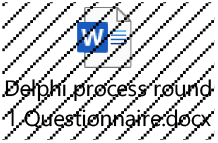

## Supplemental Appendix 2

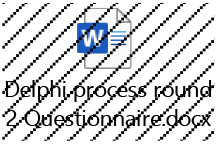

## Notes

### Competing Interest Statement

The authors have declared no competing interest.

### Funding Statement

The study was supported by funding from Wellcome Institutional Strategic Support Fund grant no. 204928/Z/16/Z through the London School of Hygiene and Tropical Medicine.

